# A population-level analysis of armed conflict and diphtheria at the subnational level in the WHO African Region 2017-2024

**DOI:** 10.1101/2024.12.18.24319262

**Authors:** Tierney O’Sullivan, Lindsay T. Keegan

**Author notes:** Author emails 1. 2.

## Abstract

**Background:** Diphtheria has been re-emerging around the world at alarming rates, raising concerns about emergency preparedness when global supplies of life-saving diphtheria antitoxin are insufficient. Outbreaks have occurred in areas with suboptimal coverage of the three-dose diphtheria tetanus and pertussis (DTP3) vaccine and regions experiencing conflict, but systematic studies assessing the association between these variables and the risk of diphtheria emergence are limited. This population-level study aimed to investigate the relationship between fatalities from armed conflict events, childhood DTP3 vaccination coverage, and the presence of reported diphtheria cases in countries in the World Health Organization’s (WHO) African region from 2017-2024.

**Methods:** The analysis was conducted at the subnational geographic scale of administrative level 1 (ADM1) (N countries=35, N ADM1 regions=541) from March 2017 to March 2024. We first used a univariate logistic regression model to establish a crude relationship between the ADM1 diphtheria status from 2017-2024 and the population-adjusted cumulative conflict-related fatalities from 2013-2024. We then fit three competing generalized logistic models with random effects accounting for weekly repeated measures at the ADM1 and country levels to estimate the relationship between time-varying rates of conflict-related fatalities and diphtheria status, adjusting for diphtheria vaccine coverage estimates.

**Results:** Results from the crude model indicate that higher ten-year cumulative rates of conflict-related fatalities are associated with a higher risk of reported diphtheria cases (OR = 1.41, 95% CI: 1.17-1.68). The results from the best-fitting repeated measures model indicate that higher rates of log-transformed conflict-related fatalities are associated with a 17.6-fold increase in diphtheria risk (OR = 17.6, 95% CI: 13.99-22.08), though risk varied widely by state and country. The best-fit model also associated lower estimates of diphtheria risk in areas with high (>80%) and low (<50%) vaccine coverage, though this is possibly due to underreporting of the true burden of disease in low-resource settings.

**Conclusions:** This exploratory analysis indicates that a history of fatalities from armed conflict is a potentially helpful indicator of subnational diphtheria risk in countries in the WHO African region from 2017-2024. Further, it may be especially useful if estimates of population-level diphtheria immunity are limited.

## Background

Diphtheria is a severe disease most commonly caused by toxigenic strains of the *Corynebacterium diphtheriae* bacterium. While it was once a leading cause of childhood morbidity and mortality (1), the introduction of effective therapeutics and the three-dose diphtheria tetanus pertussis (DTP3) vaccine have substantially reduced the burden of diphtheria cases and deaths (2). The success of campaigns such as the World Health Organization’s (WHO) Expanded Programme on Immunization (EPI) (1,2) effectively reduced diphtheria annual incidence to a low of around 5,000 cases worldwide in the mid-2010s (3). However, despite this historical progress towards control and elimination, diphtheria has been recently re-emerging at an alarming rate (3), though its re-emergence has not been homogenous (4,5).

Treatment of diphtheria requires timely administration of both antibiotics and diphtheria antitoxin (DAT): antibiotics are needed to clear the infection, while DAT is necessary to limit morbidity and mortality (6). With the success of vaccination campaigns worldwide, infectious disease specialists and public health officials were optimistic that these efforts would successfully eradicate the risk of large diphtheria outbreaks (7,8). However, due to this successful reduction in incidence, demand for DAT fell in the second half of the 20^th^ century, leading to decreased production and dwindling stockpiles, resulting in a global shortage of DAT supply and minimal manufacturing capability. Consequently, it is difficult for public health agencies to maintain or establish regional DAT stockpiles (9). Given the rapid reduction in DAT effectiveness associated with delay in treatment (10), regional DAT stockpiles are essential for timely administration to prevent fatalities from diphtheria infection (9).

Vaccination coverage and prompt administration of DAT both impact morbidity and mortality. As such, the observed case fatality rates (CFRs) among diphtheria outbreaks can vary considerably from 0-69% (1,10,11). While the average CFR declined to 7% in the 1940s-1950s, the CFR in modern outbreaks has ranged widely from 0.6–69% (10,12–14), largely because modern outbreaks have typically occurred in resource-limited settings with variable DTP3 vaccination coverage and variable access to DAT (12).

The resurgence of diphtheria in the last quarter century has often been associated with conflict, civil unrest, or population displacement: a decade-long, multi-country outbreak in the 1990s in states formed by the collapse of the Soviet Union (15), in 2017 in Bangladesh among Rohingya refugees and Yemen (13,16), and a multi-country outbreak in western Africa originating in Nigeria in 2023 (14). Understanding the regional risk of diphtheria outbreaks is important to shore up regional stockpiles of DAT to ensure prompt delivery of DAT and other public health measures to reduce infections and fatality rates.

Recently, it has been proposed that conflict and political unrest may precede diphtheria outbreaks (17). Truelove et al. (10) noted that recent outbreaks in Venezuela, Yemen, and among the Rohingya were associated with displaced populations and infrastructure failures. Dureab et al. (17) found that the risk of a diphtheria outbreak in a health district in Yemen increased by 11-fold if the district was currently experiencing conflict and that high levels of DTP3 coverage were not significantly protective when accounting for conflict. Diphtheria is a vaccine-preventable disease (VPD), and outbreak risk is highly correlated with vaccination coverage However, it is unknown what levels of DTP3 coverage equate to increased diphtheria outbreak risk or how DTP3 coverage varies with armed conflict. To investigate these relationships, we used the WHO’s strategic framework for vaccine-preventable diseases, which includes three main steps to prevent outbreaks: 1) promoting vaccine coverage, 2) adequate surveillance of emerging cases and vaccine coverage rates, and 3) emergency preparedness for large outbreaks (19). As such, we hypothesize that conflict could be related to diphtheria re-emergence risk in two ways: 1) by impacting the health infrastructure in a way that reduces vaccine coverage in the population or 2) conflict could affect public health infrastructure by reducing capacity for case surveillance or public health emergency preparedness (**Figure 1**).

**Figure 1.**
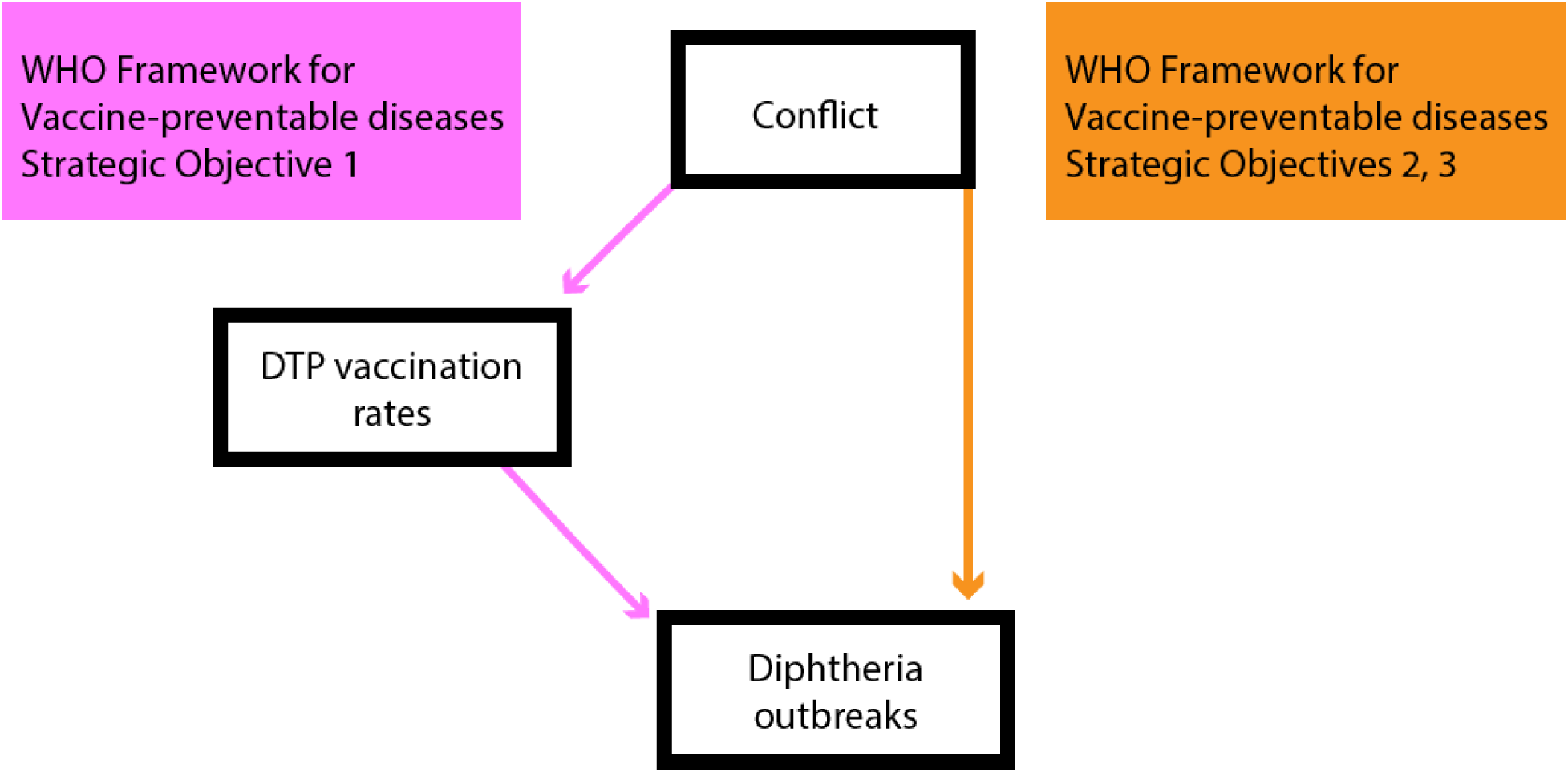
Directed acyclic graph (DAG) evaluating potential causal relationships between conflict, DTP vaccination rates, and diphtheria outbreaks. The left arm of the DAG reflects how armed conflict may affect diphtheria outbreak risk via the WHO VPD Strategic Objective 1, hypothesizing that conflict events may reduce vaccination rates, subsequently increasing the population’s susceptibility to diphtheria outbreaks. The right arm of the DAG indicates that conflict events might impact the risk of diphtheria outbreaks in a mechanism independent of reducing vaccination rates, such as population migration and crowding, or affecting the ability of areas to achieve the WHO’s VPD Strategic Objectives 2 and 3, which focus on disease surveillance and emergency response capacity.

Here, we aim to address gaps in knowledge surrounding diphtheria re-emergence and these potential risk factors by systematically investigating the relationship between diphtheria disease occurrence, DTP3 vaccination coverage, and the weekly prevalence and severity of armed conflict in the member countries of the WHO’s African region from 2017-2024. We focus our analysis on the WHO African region due to the series of recent diphtheria outbreaks occurring in the region (14). Because of high levels of within- and between-country heterogeneity of DTP3 vaccine coverage (20), armed conflict (21), and diphtheria cases throughout the WHO African region (14), we conducted our multi-country population-level analysis at a subnational geographic scale (administrative level 1 [ADM1]). This spatial scale was selected to use data at the finest spatial scale available for DTP3 vaccination coverage across the WHO African region’s member countries since country-level analyses are likely to obscure spatial clustering of unvaccinated populations that are critical for infectious disease outbreaks to arise (22).

## Methods

### Data

Since the outcome of interest is the diphtheria status of each ADM1 region over time, we used subnational diphtheria case data from the WHO African region’s weekly bulletin on outbreaks and other emergencies (23). The reports included in our analysis were published weekly from March 2017-March 2024 with rare exceptions (*see supplemental materials section S2*) and include case counts and other metadata. Although the WHO bulletins are published weekly, there were often delays in diphtheria case reporting from ongoing outbreaks, which were not updated for each location at regular time intervals. As a result of this uncertainty regarding the exact timing of reported diphtheria cases and because most diphtheria infections are asymptomatic or paucisymptomatic, it is, therefore, likely that reported cases represent an underestimate of the true burden of diphtheria infections. Consequently, we used a binary outcome of diphtheria status rather than modeling the case counts themselves. ADM1 regions were classified as being in a “diphtheria present” state if they reported more than one new diphtheria case in the past 24 weeks and “diphtheria absent” if not, even though it is possible that diphtheria transmission was occurring under the radar of detection. Both suspected and confirmed diphtheria cases were included in this case definition. For the Nigerian diphtheria outbreak, which was the largest outbreak during the study period, the WHO weekly bulletins began reporting cases in aggregate. Thus, when available, we supplemented the diphtheria case data from the more detailed Nigerian Center for Disease Control’s (NCDC) situation reports (24).

Conflict data were taken from the Armed Conflict Location and Event Data Project (ACLED) database (25). Conflict event locations were assigned to ADM1 by establishing a 1 km buffer around each latitude and longitude point location and were assigned to any ADM1 region that overlapped the buffered area. To measure conflict severity, the specific variable of interest was the total number of ACLED-reported conflict-related fatalities reported within each ADM1 in the previous 4-year period, calculated as a rate per 100,000 residents of each ADM1. Because the years of analysis were 2017-2024, the years of conflict data included in the cumulative 4-year fatalities ranged from 2013-2024.

All population-adjusted variables were established by estimating the population sizes for each ADM1 using LandScan’s 2022 global 1 km population raster (26) and overlaying with administrative polygon data from the Global Database of Administrative Regions (27).

Childhood diphtheria vaccine coverage for each ADM1 region was established from the Demographic Health Survey (DHS) variable of the estimated DPT3 vaccine coverage among children ages 12-23 months within each region (26). Since DHS surveys were not recorded annually for each country, the time-varying diphtheria coverage was estimated based on the most recent survey year until new estimates were reported.

### Inclusion criteria

Countries were eligible for inclusion if they were members of the WHO African region, had ADM1-level estimates of childhood DPT3 vaccination rates from the DHS since 2004, and had armed conflict data reported by the ACLED from 2013-2024. If a country was included, all ADM1s for that country were included in the analysis as the unit of observation (Figure 2).

**Figure 2.**
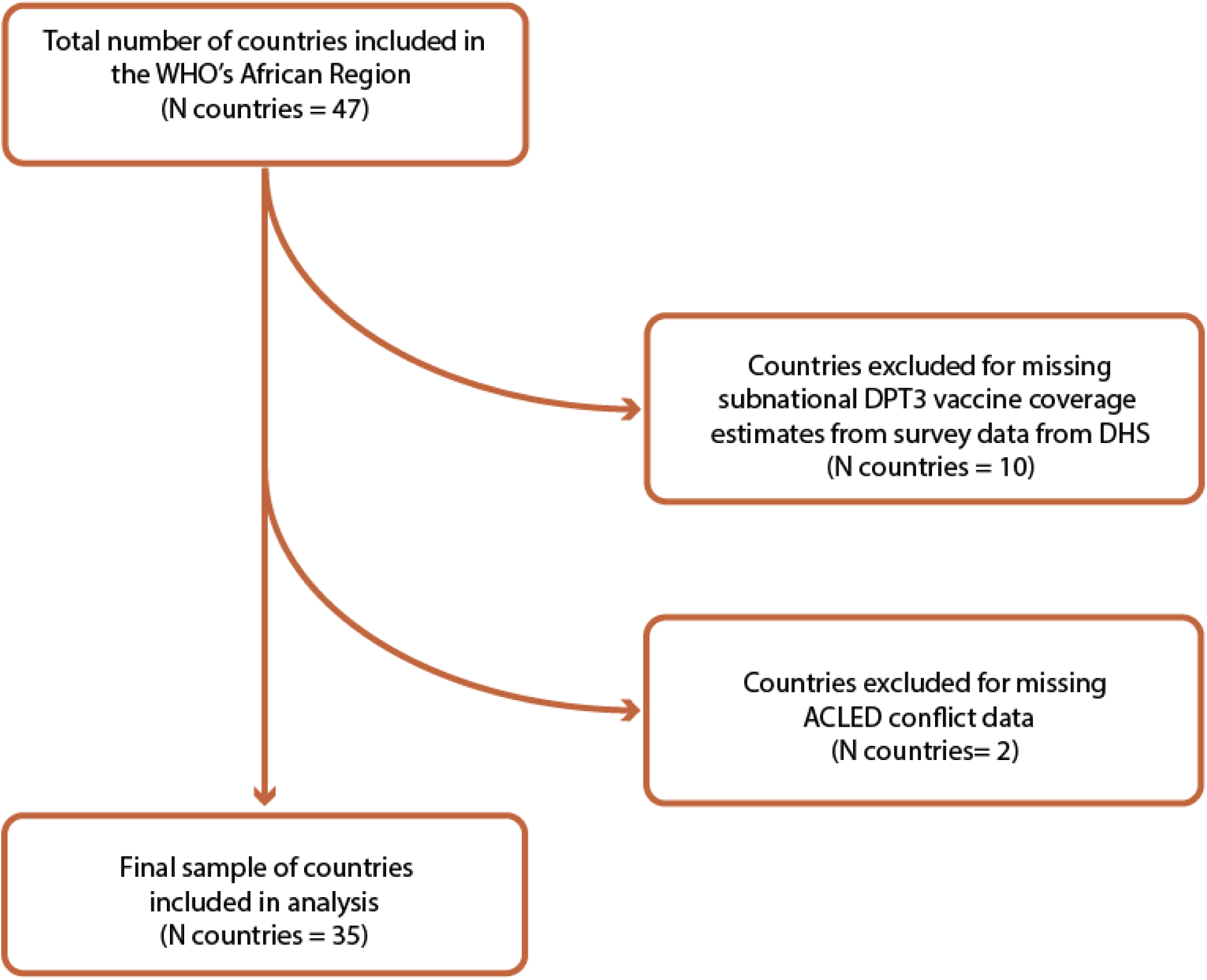
Inclusion and exclusion criteria for countries.

### Statistical analyses

All statistical analyses were conducted using the R statistical software (28).

#### Crude model

To test the crude relationship between conflict-related fatalities and diphtheria status, we first used a univariate generalized linear model with a logit link with whether diphtheria presence was ever reported from 2013-2017 as the binomial outcome and the log-transformed number of cumulative conflict-related fatalities per 100,000 residents from 2013-2024 as the sole predictor. We refer to this as the “crude model.”

#### Repeated measures models

To measure the longitudinal relationship between these conflict and diphtheria status variables and adjust for vaccine coverage, we conducted three competing mixed effects generalized linear models with logit link using the *lme4* R package (29). These “repeated measures models” include time-varying data updating at a weekly timescale and have time-varying diphtheria status (diphtheria present or absent) as the response variable. The first model, *RMCV-L* (repeated measures conflict vaccination-linear), included two linear terms as predictors: the log-transformed prior 4-year window of cumulative fatalities per 100,000 residents as the measure of conflict severity and the most recent DHS diphtheria childhood vaccine coverage estimates. The second model, *RMCV-Q* (repeated measures conflict vaccination-quadratic), included the same linear term for the measure of conflict severity and a quadratic term for vaccination coverage to address the heteroskedasticity of errors in the *RMCV-L* model. The vaccine coverage linear and quadratic terms were centered and scaled to aid in model convergence. The third repeated measures model, *RMCV-C* (repeated measures conflict vaccination-categorical), converted the vaccination coverage term from a continuous measure to a categorical with three levels of vaccine coverage: ≤50% coverage as “Low,” 50-80% as “Medium,” and ≥80% as “High,” as these categories match the existing literature’s classification of DTP3 vaccination coverage levels (20). In all three repeated measures models, we also included ADM1 (state) and ADM0 (country) as random effects, so each ADM1 and ADM0 had its own random intercept. Model comparison between the three repeated measures models was evaluated based on AIC, and where odds ratios are reported, their 95% confidence intervals (CIs) are calculated via the Wald method.

## Results

All member countries of the WHO African region (N= 47 countries) were eligible for inclusion in this analysis. Ten countries were excluded due to not having subnational data on diphtheria vaccine coverage, and two were further excluded due to not having complete conflict data, leaving 35 countries in the analysis and a total number of 541 distinct ADM1s (**Figure 2**). All ADM1 regions from the countries included in the study were included, with one exception being an ADM1 region in Mali with missing DHS survey data. See Supplementary Table 1 for a complete list of all included countries and details about the missing ADM1 region.

Overall, 47 (8.69%) of the 541 ADM1 regions had a diphtheria present status at least once in the study period. The median population-adjusted rates of cumulative conflict-related fatalities were higher among regions with diphtheria present. The median number of cumulative conflict-related fatalities per 100,000 residents from 2013-2024 in areas with diphtheria present was 9.6 (interquartile range, IQR: 3.7-16.3). In contrast, the median number of cumulative conflict-related fatalities per 100,000 residents in areas with only diphtheria-absent status was 2.6 (IQR: 1.0-7.6) (Figure 3A). The median time-weighted average of the survey-estimated childhood vaccination coverage was higher in areas that never reported the presence of diphtheria cases (median: 80.6, IQR: 67.7-89.0) than in ones that reported diphtheria presence at least once (median: 66.2, IQR: 35.6-71.1) (Figure 3B). The number of ADM1 regions that reported diphtheria presence also increased over time, with the vast majority occurring in 2023-2024 (Figure 4).

**Figure 3.**
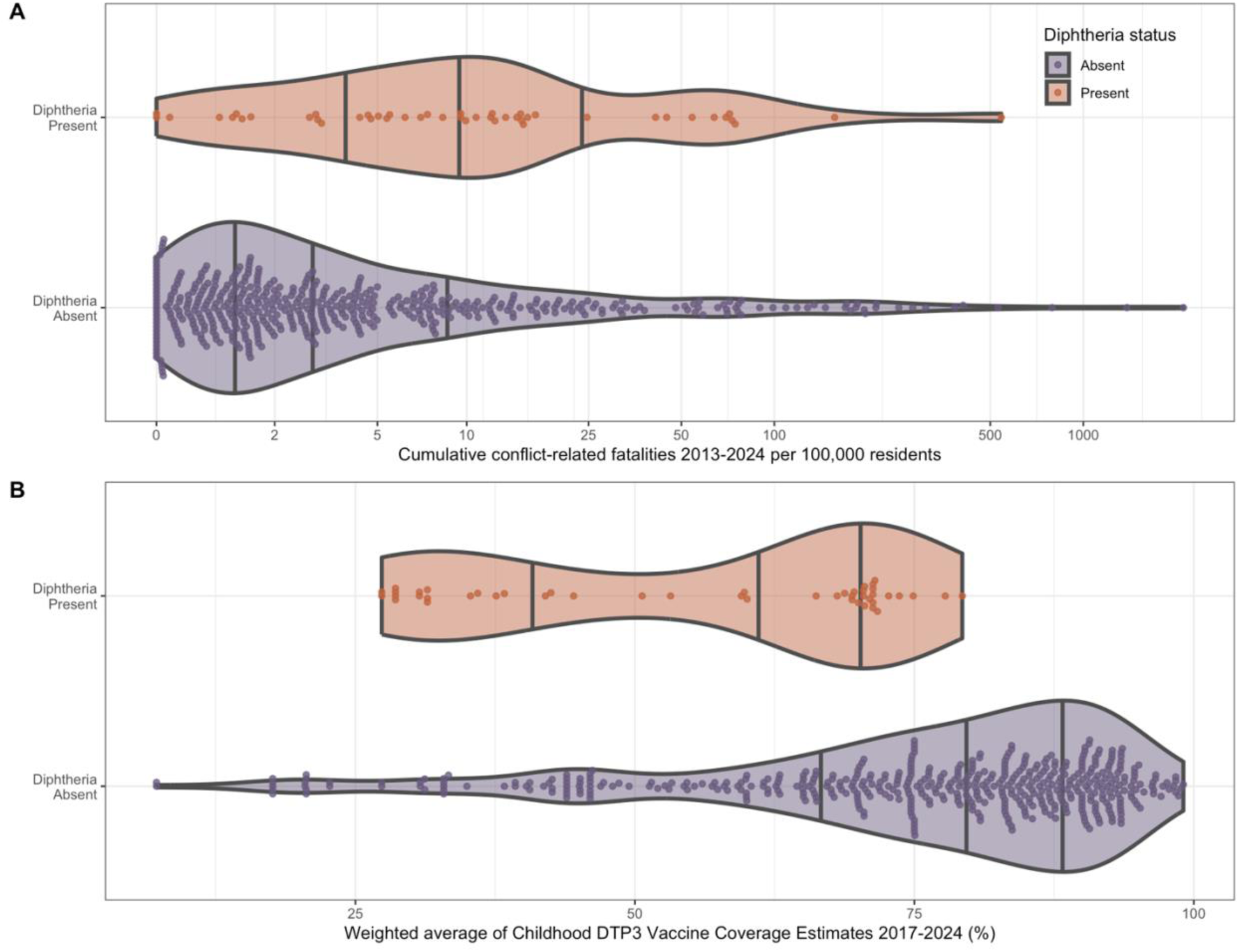
Violin plots depicting the densities of observed data for each predictor variable by the diphtheria status of each administrative level 1 (ADM1) region. “Diphtheria present” regions reported diphtheria cases at least once during 2017-2024 (ADM1 regions, N = 47), whereas “diphtheria absent” regions were never classified as diphtheria present during the study period (ADM1 regions, N = 494). The vertical gray lines within each density plot indicate the minimum, 25^th^ quartile, median, 75^th^ quartile, and maximum values, respectively. A) The cumulative population-adjusted counts of conflict-related fatalities from 2013-2024 for each administrative level 1 (ADM1) region in the analysis, with the x-axis on a log scale for readability. B) The time-weighted average of survey-estimated childhood three-dose diphtheria-tetanus-pertussis vaccination coverage for each ADM1 region from 2017-2024.

**Figure 4.**
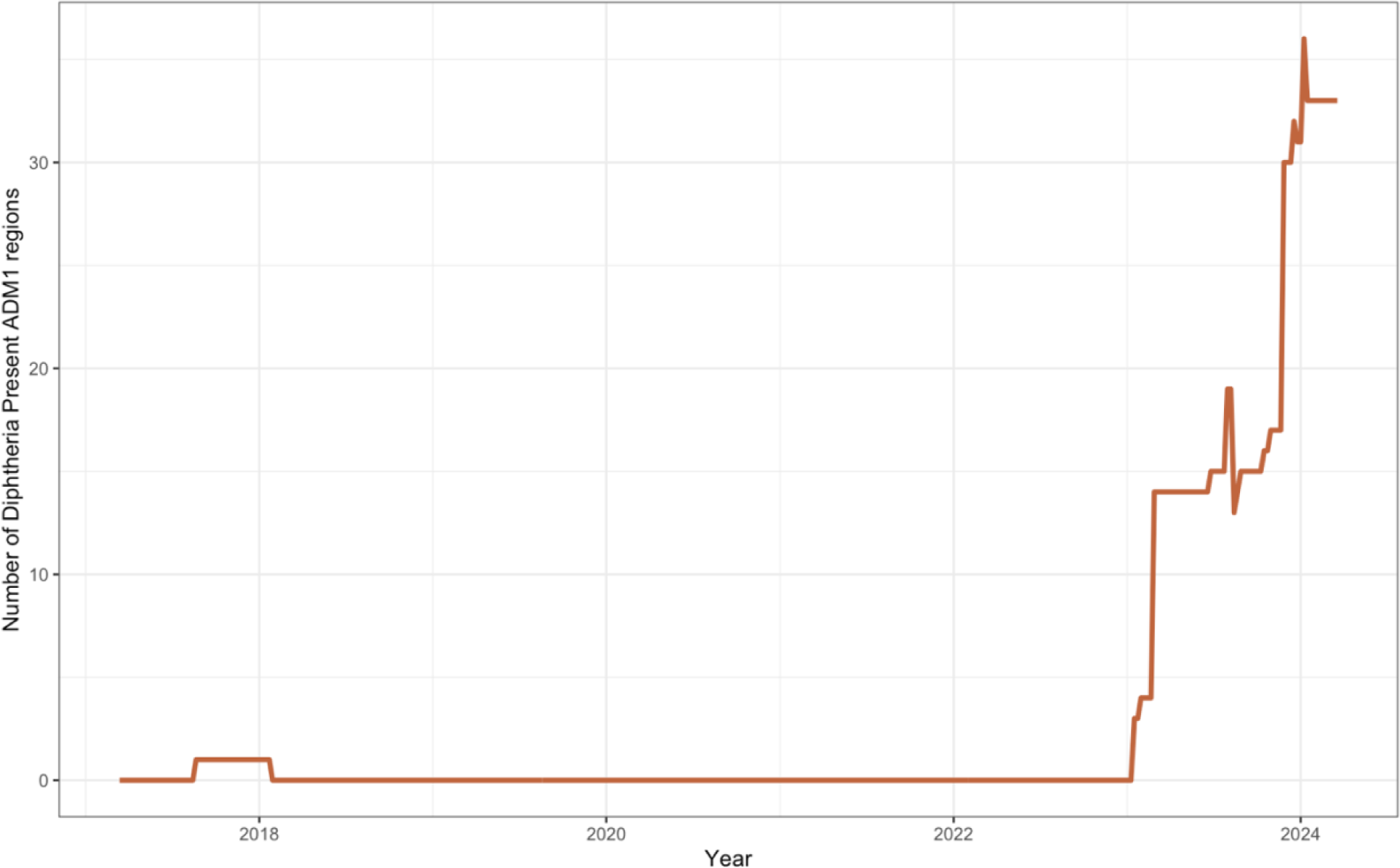
The number of ADM1 regions in the analysis classified as “diphtheria present” over the study period from March 2017 to March 2024.

In the crude model, which only assessed the relationship between total conflict-related fatalities from 2013-2024 and whether each ADM1 region ever experienced a diphtheria present status from 2017-2024, the crude odds ratio between the relationship of conflict-related fatalities and whether the ADM1 region ever reported diphtheria present is 1.41 (95% CI: 1.17-1.68, *p < 0.001*). This indicates that without accounting for temporality or spatial dependence, with one increase in the unit of the logarithm of cumulative conflict-fatalities, the probability of reporting the presence of diphtheria increased by 41% (Table 1).

**Table 1.**
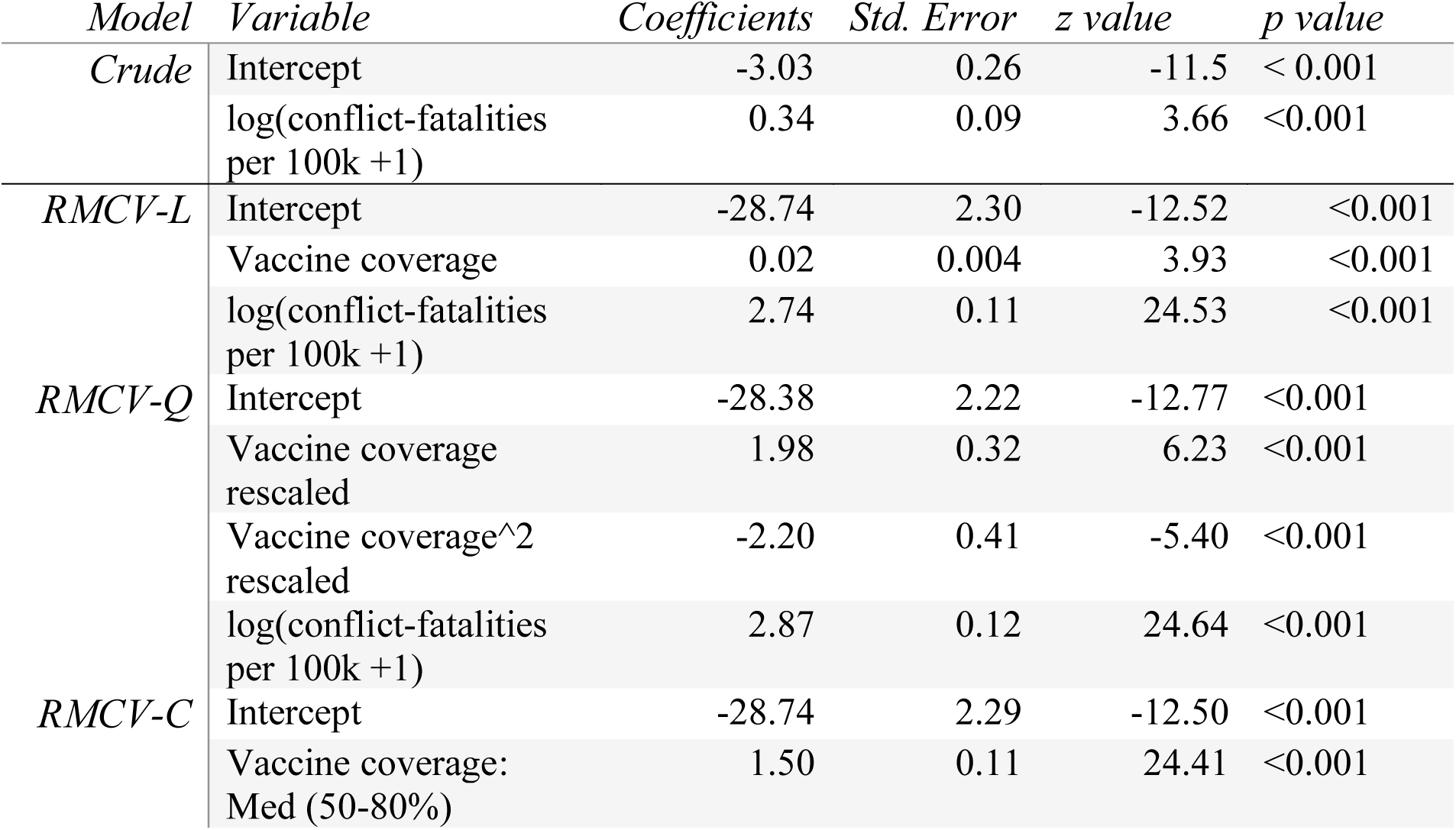
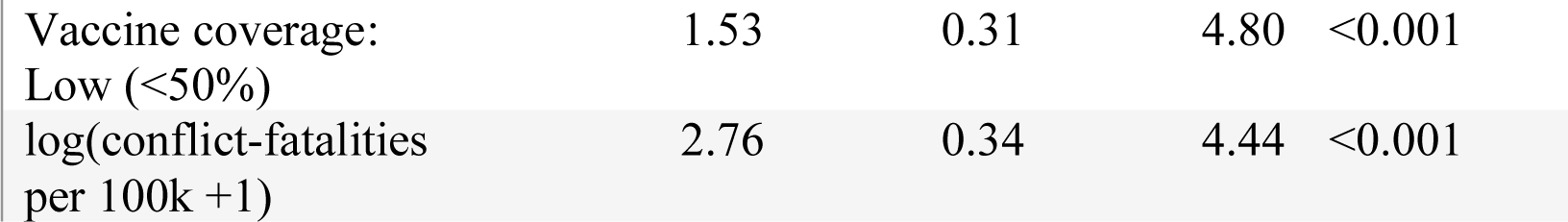
Model-estimated coefficients and their standard errors.

| *Model* | *Variable* | *Coefficients* | *Std. Error* | *z value* | *p value* |
| --- | --- | --- | --- | --- | --- |
| *Crude* | Intercept | -3.03 | 0.26 | -11.5 | < 0.001 |
|  | log(conflict-fatalities per 100k +1) | 0.34 | 0.09 | 3.66 | <0.001 |
| *RMCV-L* | Intercept | -28.74 | 2.30 | -12.52 | <0.001 |
|  | Vaccine coverage | 0.02 | 0.004 | 3.93 | <0.001 |
|  | log(conflict-fatalities per 100k +1) | 2.74 | 0.11 | 24.53 | <0.001 |
| *RMCV-Q* | Intercept | -28.38 | 2.22 | -12.77 | <0.001 |
|  | Vaccine coverage rescaled | 1.98 | 0.32 | 6.23 | <0.001 |
|  | Vaccine coverage^2 rescaled | -2.20 | 0.41 | -5.40 | <0.001 |
|  | log(conflict-fatalities per 100k +1) | 2.87 | 0.12 | 24.64 | <0.001 |
| *RMCV-C* | Intercept | -28.74 | 2.29 | -12.50 | <0.001 |
|  | Vaccine coverage: Med (50-80%) | 1.50 | 0.11 | 24.41 | <0.001 |

|  |  |  |  |  |
| --- | --- | --- | --- | --- |
| Vaccine coverage:<br>Low (<50%) | 1.53 | 0.31 | 4.80 | <0.001 |
| log(conflict-fatalities<br>per 100k +1) | 2.76 | 0.34 | 4.44 | <0.001 |

The best-fitting model of the three repeated measures models was the *RMCV-Q* model, which included both linear and quadratic terms for childhood DTP3 vaccination coverage (ΔAIC = 18.92). All three repeated measures models accounted for temporality between the conflict-related fatalities, childhood DTP3 vaccine coverage estimates, and ADM1 diphtheria status while also accounting for random effects for ADM1 and ADM0 locations. Compared to the crude model, the odds ratio for conflict severity in the repeated measures models increased substantially to 16-18. In the best fitting model, *RMCV-Q*, the odds ratio for the conflict-related fatalities was 17.6 (95% CI: 13.99-22.08, *p* < 0.001), indicating that an increase in the log number of population-adjusted 4-year conflict-related fatalities is associated with a 17.6 times higher risk of reporting the presence of diphtheria cases. Though the model-predicted probability of diphtheria risk increased with higher conflict severity, the predicted risk estimates varied depending on the random intercepts for AMD0 and ADM1 (Figure 5).

**Figure 5.**
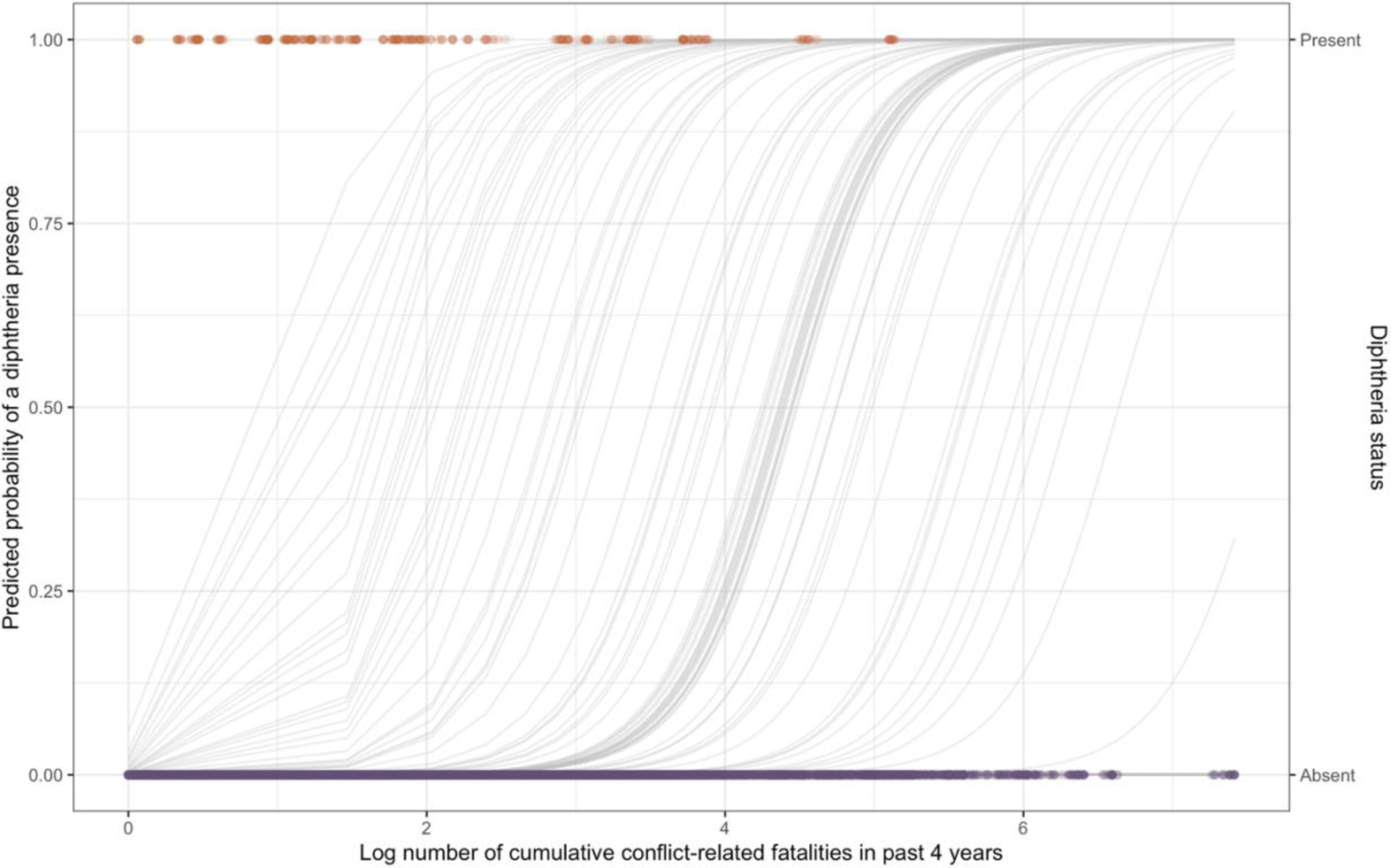
The predicted probability of reporting diphtheria presence (grey lines) for each administrative level 1 (ADM1) by the log-transformed number of conflict-related fatalities in the previous 4 years. Lines are shifted left or right depending on their random intercepts for each country and ADM1. Orange points along the top indicate the observed data indicating ADM1 regions with reported diphtheria presence. Purple points along the bottom of the graph indicate observed ADM1 regions with diphtheria absent status. To illustrate a single model-predicted probability of diphtheria presence for each ADM1 region, vaccination rates were set to the median for each location.

The repeated measures models also included survey-based estimates of childhood DTP3 vaccination coverage. Surprisingly, in the *RMCV-L* model, DTP3 vaccination rates were minimally associated with diphtheria status (OR = 1.02, 95% CI: 1.01-1.03, *p* <0.001). The direction of this relationship was the opposite of what was expected, as it indicates that a one percent increase in childhood vaccination coverage is associated with a 2% increase in the risk of a diphtheria presence. This model was outperformed by the *RMCV-Q* model (ΔAIC = 28.48), indicating that the linear fit of vaccine coverage to diphtheria presence was not appropriate.

The *RMCV-Q* model included an additional quadratic term for DTP3 vaccination coverage, which helped address some heteroskedasticity of the errors in the *RMCV-L* model. This changed the model-estimated relationship between vaccination coverage and diphtheria presence, where diphtheria presence risk was lowest in areas with low (<50 %) and high (>80%) DTP3 vaccine coverage.

The final repeated measures model, *RMCV-C,* included vaccination coverage as a categorical term to the model, with levels of low (≥50%), medium (50-80%), and high (>80%). With the high level of vaccination as the reference, the model estimated odds ratios for medium vaccination coverage was 4.48 (95% CI: 2.42-8.26, *p < 0.001*), and low vaccination coverage was 4.60 (95% CI: 2.35-9.03, *p < 0.001*). This indicates that having high vaccination coverage of 80% or more was associated with a protective effect against diphtheria, with areas with medium or low coverage associated with a 4.48 and 4.60-fold increase in reporting diphtheria cases present. However, there was substantial overlap between the 95% confidence intervals for low and medium vaccine coverage. This model was outperformed by the RMCV-Q model (ΔAIC = 18.92), indicating that the quadratic fit for vaccination was more optimal than using categorical vaccination levels.

## Discussion

Here, we investigated the relationship between population-level risk of reported diphtheria cases and regional conflict, measured by conflict-related fatalities. Our model provides evidence supporting a strong relationship between historical conflict severity and subsequent diphtheria outbreaks, even when accounting for random effects of each state and country and when accounting for childhood vaccine coverage estimates. Our results also indicate that conflict-related fatalities are a better predictor of subnational reported diphtheria presence than estimates of childhood DPT3 vaccine coverage alone in the WHO African region member countries. This supports similar findings from a subnational study of conflict and diphtheria in Yemen in 2017 (18). Surprisingly, we found that rather than a monotonically decreasing relationship between DTP3 childhood vaccination rates and the risk of diphtheria presence, the relationship determined from our best-fitting model was quadratic with a peak in reported diphtheria presence among states with DTP3 between 50% and 80%. One potential explanation for this observed relationship is due to a high degree of misclassification of childhood DTP3 coverage in our analysis, which was based on surveys conducted at irregular time intervals by the DHS and not on systematic reporting of vaccination administration directly (26). Another possible rationale is that childhood DTP3 vaccination coverage does not adequately represent the overall population level of immunity against diphtheria, either from historical childhood DTP3 vaccination rates, partial childhood vaccination (i.e., DTP1), adult booster coverage, or immunity from prior infection (30). A third explanation may be that places with low vaccination coverage have poor health infrastructure and may not have the surveillance systems to detect diphtheria cases. In contrast, places with high vaccination coverage may have surpassed the critical vaccination threshold, preventing diphtheria spread. Seroprevalence surveys, along with fine-scale DTP3 and booster vaccination coverage data, may help illuminate the observed limited impact of DTP3 vaccination rates on diphtheria outbreak risk (31). Although these are costly and unlikely to be conducted at scale or regular intervals, given the substantial increase in diphtheria presence following the COVID-19 pandemic, a deeper understanding of diphtheria risk is crucial.

Our results indicate that data on recent armed conflict may be helpful for public health response planning, particularly in areas with limited access to vaccination coverage data. The high degree of dangerous and violent conflict events may limit the usefulness of this tool in affected regions since efforts to bolster public health infrastructure may not be feasible in these highest-risk locations (32). Even if this is the case, having insights into the risk of diphtheria outbreaks in these populations could still guide regional resource planning for diphtheria antitoxin stockpiles, training clinicians to promptly recognize diphtheria symptoms, or establish laboratory capacity for expedited confirmatory testing. This information is relevant to guide planning in geographic regions surrounding conflict-affected areas, especially if there are large migrations of individuals from high diphtheria risk areas as refugees (13).

There are a number of limitations of this analysis. The data of reported diphtheria cases are likely an underestimate of the true burden of disease. However, it is expected that in areas with destabilized public health infrastructure due to higher intensity and frequency of conflict events, the surveillance would be less effective, decreasing the probability of detecting diphtheria cases and small outbreaks. Thus, we expect that if all diphtheria cases were accurately reported, this would strengthen the observed relationship between past conflict and diphtheria risk rather than mitigating it. Another limitation is that this analysis does not take into account the outbreaks of neighboring countries or temporal autocorrelation. Future studies could expand on this exploratory analysis to establish more robust estimates of the risk factors for diphtheria outbreaks by including a mechanism for diphtheria case importation and a model incorporating a time-dependent error structure.

## Conclusions

We found that a local history of severe armed conflict, as assessed by the number of resulting fatalities, is associated with subsequent reports of diphtheria presence in Africa from 2017-2024 and should be considered as a potential early signal of increased outbreak risk. Although high levels of childhood DTP3 vaccine coverage were protective against the presence of reported diphtheria cases, we found that the relationship was somewhat complex, with diphtheria risk peaking between 50-80% DTP3 coverage. However, this may be an artifact of low diphtheria case reporting in low vaccine coverage areas. Because of this, we suggest that the history and severity of armed conflict may be an early indicator of increased risk of diphtheria if local vaccination coverage data are unavailable, as is often the case in low-resource settings. Even for areas with reliable historical DTP3 vaccination coverage data, the ACLED armed conflict data are particularly useful due to their real-time reporting of geolocated conflict events. They are also available more quickly and at a finer spatial scale than most vaccination coverage estimates. As conflict increases in intensity and frequency, and the number of refugees and internally displaced people (IDP) has increased across the globe (33), this information may become more salient for public health agencies to prepare for re-emerging diphtheria outbreaks and infectious disease emergencies in general (34).

## Data Availability

Diphtheria case count data are publicly available and were originally located at the WHO African Region's Weekly Bulletins of Outbreaks and other Emergencies.
Additional diphtheria case data were obtained from the Nigerian CDC situation reports.
Data on fatalities from armed conflict events were obtained from the Armed Conflict Location and Event Database, which is publicly accessible here.
And aggregated data on state-level childhood vaccination coverage estimates were obtained from the Demographic Health Surveys.

https://dhsprogram.com/data/

https://acleddata.com/data/

https://ncdc.gov.ng/ncdc.gov.ng/diseases/sitreps/?cat=18&name=An%20Update%20of%20Diphtheria%20Outbreak%20in%20Nigeria

https://www.afro.who.int/health-topics/disease-outbreaks/outbreaks-and-other-emergencies-updates

## List of abbreviations

WHO: World Health Organization
DAT: diphtheria antitoxin
VPD: vaccine-preventable disease
DTP3: three-dose diphtheria tetanus pertussis
ADM1, ADM0: vaccine, administrative level 1, 0
DHS: Demographic Health Survey
ACLED: Armed Conflict Location and Event Data

## Supplementary Materials

### Supplement 1. (S1)

**Supplementary Table 1 (ST1).**
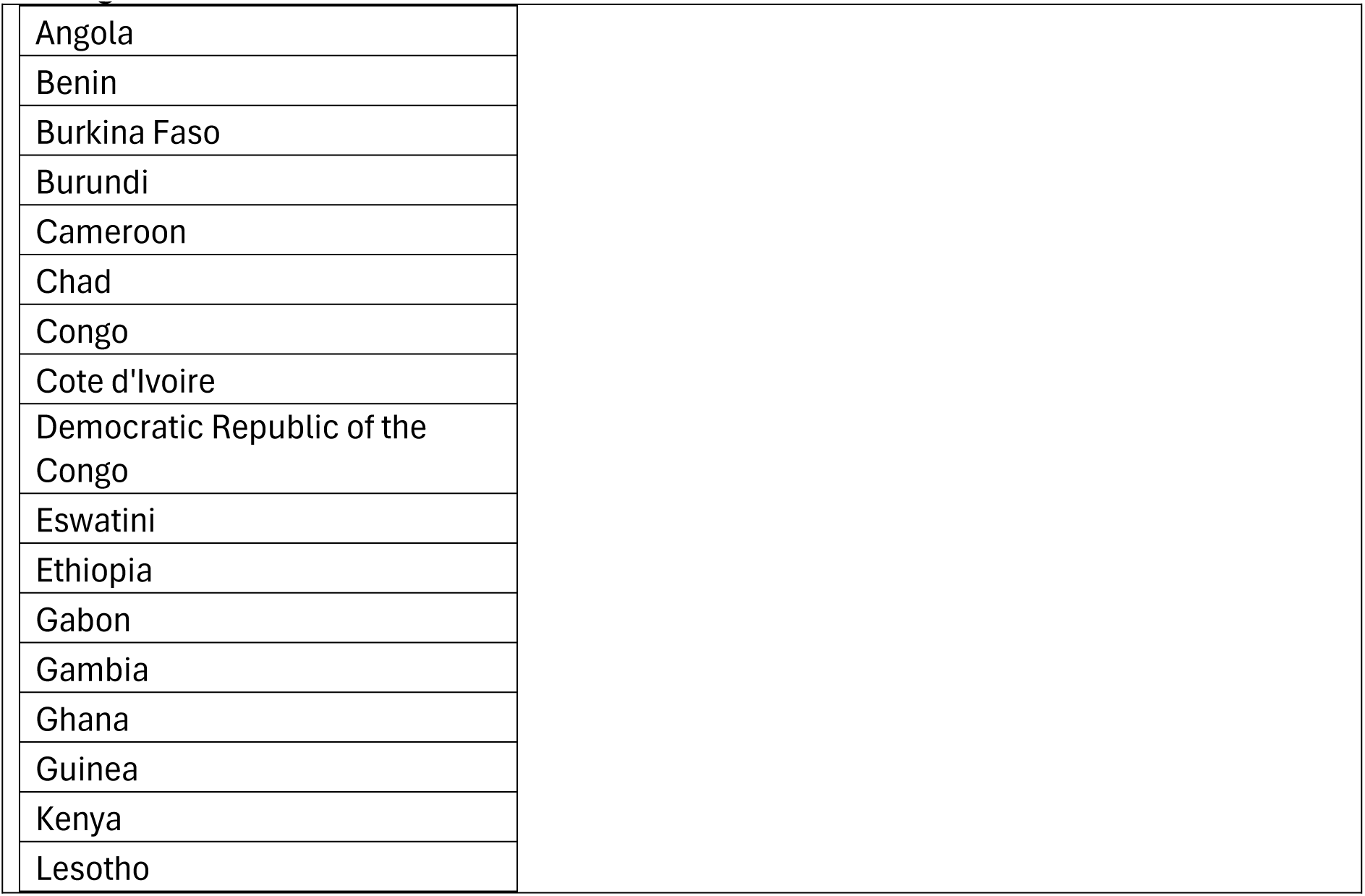
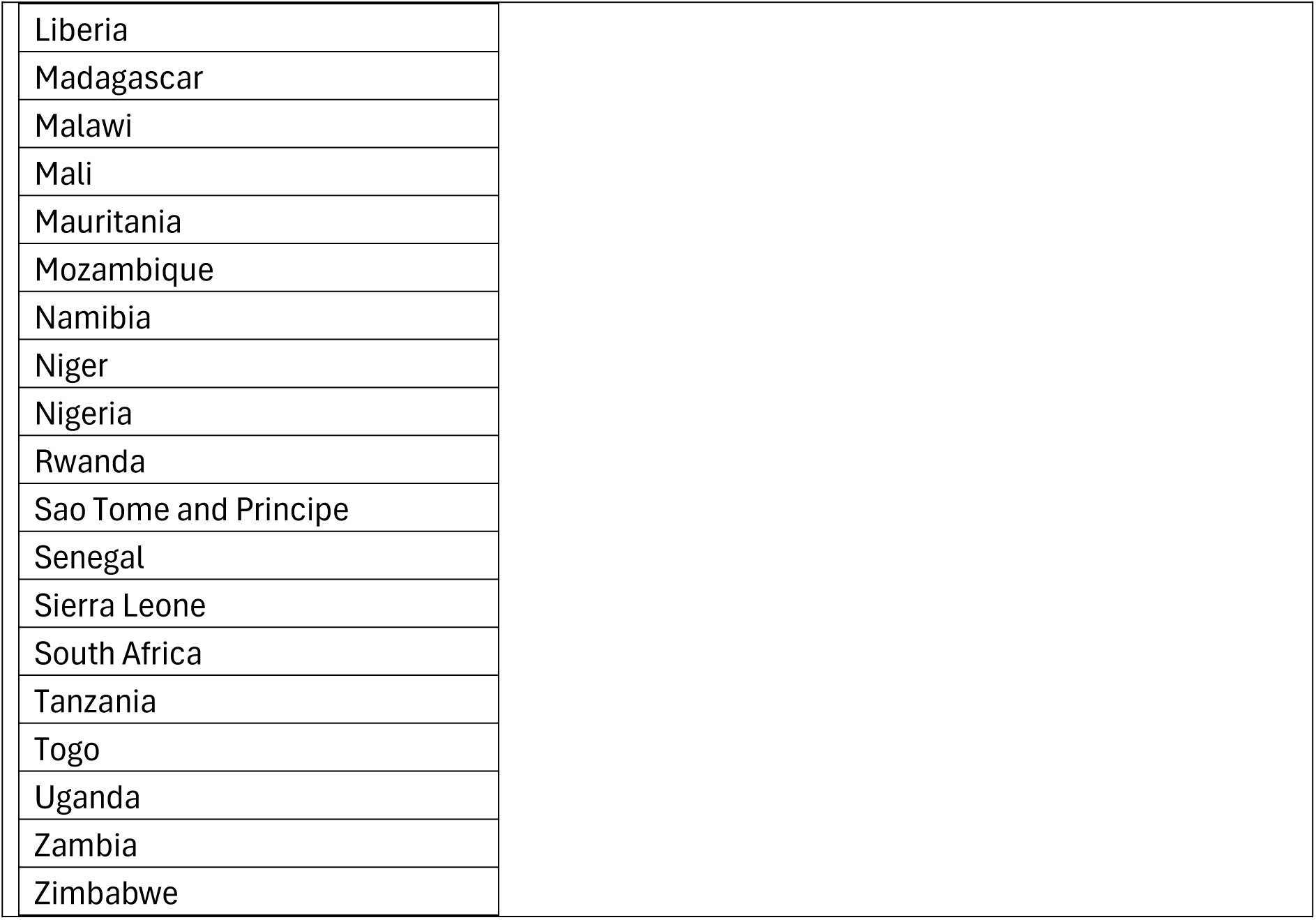
A List of countries in the WHO Africa Region included in the analysis. Of the listed countries, all ADM1 regions were included in the study except for the Kidal region in Mali, which was excluded from available DHS surveys of DTP3 vaccine coverage.

### Supplement 2 (S2)

**Missing weeks of data from WHO African Region’s weekly bulletins of outbreaks and other emergencies.**

We extracted data on timing and locations of suspected and confirmed diphtheria cases from the WHO African Region Weekly Bulletins on Outbreaks and other Emergencies from ISOWeek 10 in March of 2017 through ISOWeek 11 in March of 2024. Of the 367 weeks, 11 (3%) were missing from the online repository of weekly bulletins, which included ISOWeeks 12, 43-46, 50, and 52-53 of 2020; ISOWeek 17 of 2021, and ISOWeeks 31-32 of 2023.

